# Description of Kratom Exposure Events in Wisconsin as Reported to the Wisconsin Poison Center — January 1, 2010–September 1, 2022

**DOI:** 10.1101/2023.01.03.22284038

**Authors:** Peter DeJonge, David Gummin, Nicholas Titelbaum, Jonathan Meiman

## Abstract

**Background:** Consumption of kratom (*Mitragyna speciosa*), an herbal substance, can result in adverse health effects. We characterized kratom-associated adverse events in Wisconsin to provide pertinent recommendations for clinicians and public health practitioners.

**Methods:** Using Wisconsin Poison Center (WPC) data, we searched for and summarized all records associated with exposure to “kratom”, “electronic delivery device containing kratom”, or “mitragyna” during January 1, 2010–September 1, 2022.

**Results:** Kratom-associated exposure calls to WPC increased 3.75 times during 2016–2020. Among all 59 calls, 26 (44.1%) reported concomitant use of another substance, agitation was the most common symptom reported (23, 39%), and 7 persons required critical care. Three unintentional ingestions were reported in infants aged <2 years.

**Discussion:** Kratom-associated exposure calls to WPC have been generally increasing in frequency since 2011. Wisconsinites who choose to use kratom might benefit from education regarding health risks and safe storage practices to avoid unintentional pediatric exposure.

## BACKGROUND

Kratom is an herbal substance derived from the leaves of *Mitragyna speciosa*, a tree native to Southeast Asia, and is commonly consumed in a tea or as a dried powder.^1^ Two principal kratom alkaloids, mitragynine and 7-hydroxymitragynine, are responsible for kratom’s psychotropic properties, which range from stimulant-like effects at low doses to opioid-like sedative effects at higher doses.^2^ Kratom is often ingested for self-management of pain, anxiety, depression and to stop or reduce opioid use or alleviate withdrawal symptoms.^3^

Although considered a “drug of concern” by the U.S. Drug Enforcement Agency, kratom remains unscheduled by the U.S. Controlled Substances Act and its legality is determined on a state-by-state basis.^4^ Wisconsin is 1 of 6 states where possession of kratom is illegal statewide and thus not subject to commercial regulation.^3^ However, kratom use still occurs in Wisconsin and is therefore important to understand both clinically and from a public health perspective given the range of kratom-associated adverse events reported in literature.^1^ We examined data from the Wisconsin Poison Center (WPC) during January 1, 2010–September 1, 2022, to characterize kratom-associated adverse events in Wisconsin and provide pertinent recommendations for clinicians and public health practitioners.

## METHODS

WPC data are shared with the National Poison Data System (NPDS), a collection of data logged by all poison centers in the United States and maintained by America’s Poison Centers.^5^ We queried NPDS for all Wisconsin-originated records associated with “kratom” (generic code: 0310130, product code: 7224390), “electronic delivery device containing kratom” (product code: 8306048), or “mitragyna” (product code: 4271683). We searched all records generated during 23 January 1, 2010–September 1, 2022.

We only considered calls associated with substance exposure (i.e., calls for the purposes of drug identification or information-gathering were excluded). Kratom-exposure calls were characterized by year of exposure, county of caller, reason for call, demographic characteristics, single vs polysubstance exposure, reported symptoms, highest level of healthcare received, and overall medical outcome. These categories follow NPDS coding schemes developed by America’s Poison Centers.^5^ Fisher’s exact test was used for unadjusted comparisons of categorical variables. We also summarized narrative information from exposure calls associated with the most severe medical outcomes. R was used to complete all data analyses and figures (version 4.1).^6^ This activity was reviewed by CDC and was conducted consistent with applicable federal law and CDC policy.^†^

## RESULTS

During January 1, 2010–September 1, 2022, WPC received 59 calls associated with kratom exposure (Table 1). Most exposed persons were self-reported male (37/59, 62.7%). One person reported being pregnant at time of exposure. Of 52 (88.1%) calls with age information available, the mean age of exposed persons was 35.3 years (standard deviation = 15.4 years). Three exposures occurred among children aged <18 years; all 3 were among infants aged <2 years and reported as unintentional ingestions. Each of these 3 pediatric exposures was recorded by WPC staff as associated with little-to-no medical outcome; however, 1 infant was admitted to the pediatric intensive care unit for observation.

**Table 1.** Characteristics of All Kratom-Associated Exposure Calls (N = 59) to the Wisconsin Poison Center — January 1, 2010–September 1, 2022

| Exposure characteristics | No. (%) |
| --- | --- |
| Female | 22 (37.3) |
| Age in years, mean (sd) | 35.3 (15.4) |
| Reason for call |  |
| Adverse reaction to drug | 8 (13.6) |
| Intentional — abuse, misuse, or unclear reason | 38 (64.4) |
| Intentional — suspected suicide | 6 (10.2) |
| Withdrawal symptoms | 2 (3.4) |
| Unintentional | 3 (5.1) |
| Unknown or missing | 2 (3.4) |
| Symptom reported<sup>A</sup> |  |
| Agitation | 23 (39.0) |
| Tachycardia | 21 (35.6) |
| Confusion | 14 (23.7) |
| Central nervous system depression | 13 (22.0) |
| Hypertension | 9 (15.3) |
| Medical outcome<sup>B</sup> |  |
| No effect | 7 (11.9) |
| Mild effect | 28 (47.5) |
| Moderate effect | 16 (27.1) |
| Major effect | 3 (5.1) |
| Death | 1 (1.7) |
| Unable to assess, lost to follow-up | 4 (6.8) |
| Highest level of healthcare facility care |  |
| Unknown or refused treatment | 23 (39.0) |
| Admit, treat and release | 17 (28.8) |
| Admit, noncritical<sup>C</sup> | 12 (20.3) |
| Critical care admission | 7 (11.9) |

<sup>A</sup> Multiple symptoms were able to be reported by exposed persons. Here, the five most frequently reported symptoms are presented.
<sup>B</sup> Defined by the National Poison Data System (NPDS) as the “Medical outcome of the patient following exposure based on all available information”. *No effect* reflects a combination of two NPDS outcome categories: “No effect” and “Unrelated effect, the exposure was probably not responsible for the effect(s)”. A *minor effect* was defined as “the patient exhibited some symptoms as a result of the exposure, but they were minimally bothersome to the patient”. A *moderate effect* was defined as “the patient exhibited symptoms as a result of the exposure which are more pronounced, more prolonged or of a more systemic nature than minor symptoms”. A *major effect* was defined as “the patient exhibited symptoms as a result of the exposure which were life-threatening or resulted in significant residual disability or disfigurement”.
<sup>C</sup> Includes one exposed person who was recorded as “admitted to psychiatric facility”.

After zero calls reported in 2010, kratom exposure associated calls increased from 1 call in 2011 to a peak of 15 calls in 2020 (Figure 1); based on visual inspection there were no obvious changes over time in the patterns of medical outcome or polysubstance exposure. Among exposures with county information (N = 54), the majority were concentrated in southeastern Wisconsin counties, containing the Madison and Milwaukee metropolitan areas (Figure 2). Marinette County in northeast Wisconsin reported the highest number of kratom exposures (10, 18.5%), which were distributed over time (1 in 2018, 4 in 2019, 2 in 2020, 2 in 2021, and 1 in 50 2022).

**Figure 1.**
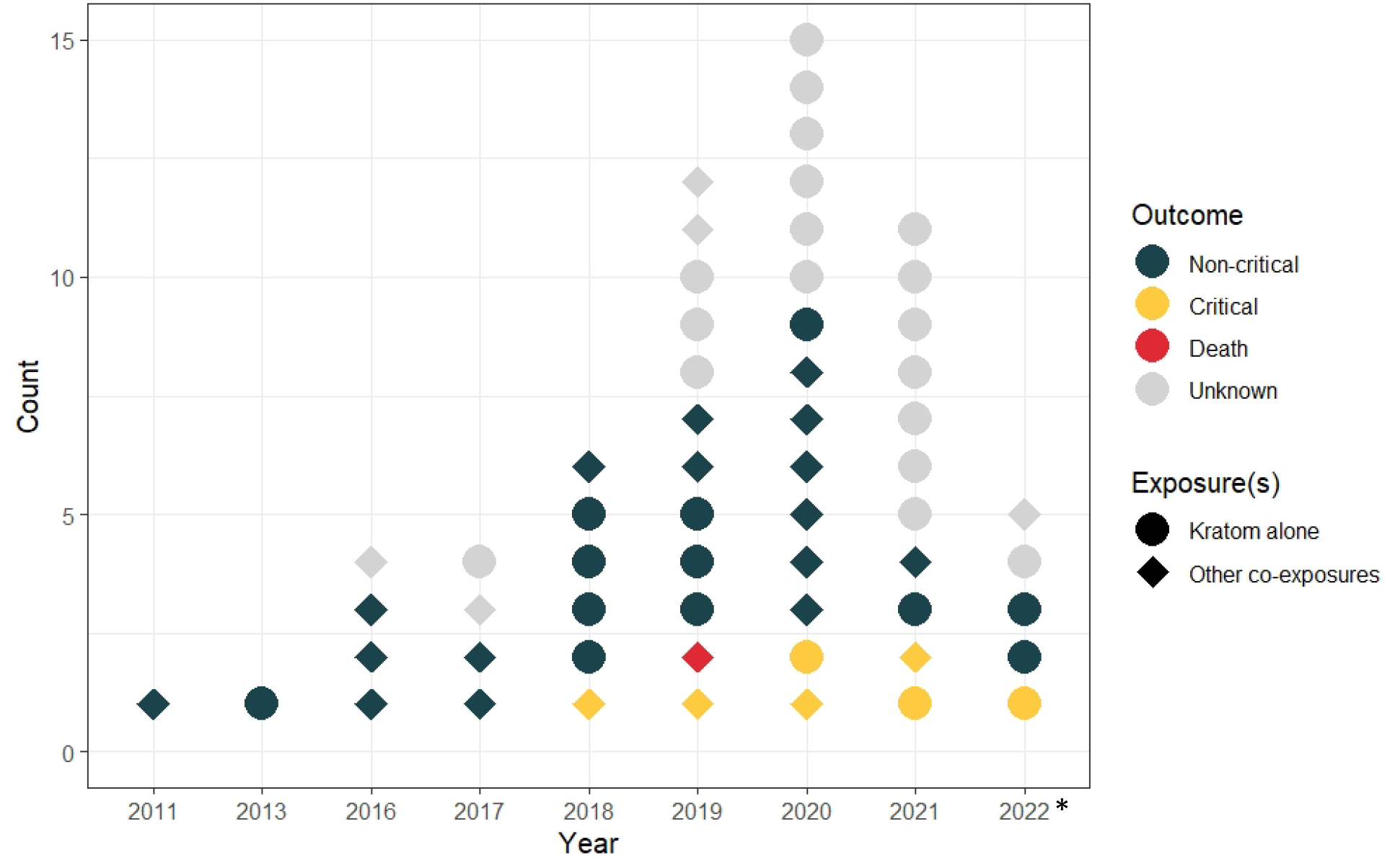
Timeline of Kratom-Associated Exposure Calls (N = 59) to the Wisconsin Poison Center — January 1, 2010–September 1, 2022 *\*The year 2022 is denoted with an asterisk given the incomplete nature of the data at time of analysis*.

**Figure 2.**
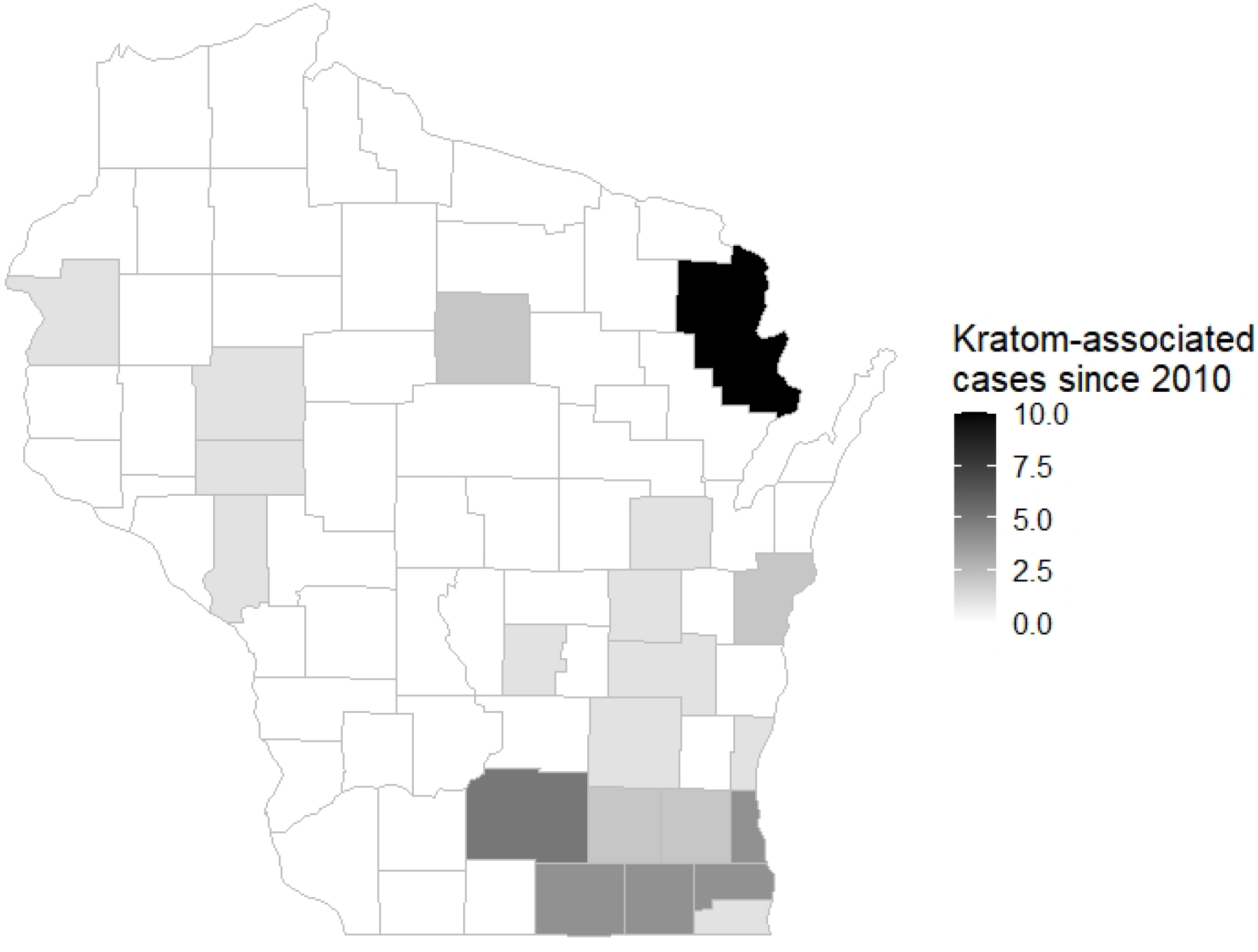
Distribution of Kratom-Associated Exposure Calls with County Information (N = 54) Recorded by the Wisconsin Poison Center — January 1, 2010–September 1, 2022

Approximately half of callers reported kratom as the only exposure substance (N = 33, 55.9%). Kratom exposure by itself, compared with polysubstance exposure, generally occurred in younger persons (mean age = 31.9 years vs 38.7 years, respectively). Among persons reporting polysubstance exposures, the most common co-substances were alcohol (N = 8, 30.8%) and benzodiazepines (N = 3, 11.5%). Fisher’s exact test for association indicated that compared with exposures of kratom alone, polysubstance exposure was not significantly associated with medical outcome reported (*P* = .22) nor level of healthcare received (*P* = 1.0), though these analyses are limited by small numbers.

Agitation (N = 23; 39%), tachycardia (21; 35.6%), confusion (14; 23.7%), and generalized central nervous system depression (13; 22.0%) were the most commonly reported clinical findings. Among 50 calls with known medical outcome, 19 (38.0%) were reported with moderate or major medical outcomes. Among 36 calls with known levels of healthcare received, critical care was required for 7 persons (22.2%) although only 1 received laboratory confirmation of kratom exposure; 5 presented with marked agitation and required sedation therapy and 3 required mechanical ventilation.

Among critical care admissions, 1 was an infant with suspected kratom exposure. The infant, presenting with tachycardia and vomiting, was kept overnight in the pediatric intensive care unit for monitoring; the child was reported normal at discharge the following day. Additionally, in different years and counties, 2 males in their early 30s were admitted to critical care. Both were active weightlifters, presented with agitation, and reported co-ingestion of phenibut, a central nervous system depressant unregulated in the United States and commonly marketed online as a dietary supplement.

WPC also recorded 2 critical care admissions among females in their 70’s. Both presented with tachycardia, confusion, and marked agitation. One of the women died in the hospital with sepsis complications, though postmortem toxicology identified kratom as contributory. During initial presentation at a local emergency department, a family member reported the patient’s recent use of kratom for chronic pain—believed to be ≥1 18 mg kratom capsule daily. A capsule source was not identified. A quantitative serum mitragynine level was obtained on hospital admission and returned at 26 ng/ml.

## DISCUSSION

In Wisconsin, kratom-associated exposure calls to WPC have been generally increasing in frequency during the past decade—similar to the trend nationwide.^7^ Though the number of studies on kratom use is increasing also, the literature still lacks a consensus as to the substance’s health benefits and risks.^8^ For one, analyses of U.S. kratom use are challenged by the limitations of passive surveillance systems,^7,9^ which likely undercounts kratom-associated adverse events. Neither traditional drug tests nor forensic toxicology assays generally screen for mitragynine.^8^ Secondly, in the absence of governmental or commercial kratom regulation, research is often unable to categorize the potency, quality, or actual substance being consumed.^10^

An additional complication in our understanding of kratom-associated outcomes is the considerable prevalence of polysubstance exposure—recorded in approximately half of WPC calls in our project. Clinicians and public health practitioners may consider cautioning people against use of kratom concomitant with other substances due to unknown possible harmful drug interactions.^2,7^ This message is perhaps particularly relevant among older adults, such as the two women in their 70’s in our analysis, who are more at risk for adverse drug interaction outcomes because of their high prevalence of prescription medication use.

Kratom use education may also consider prioritizing messaging among adults with children or expectant parents. WPC recorded 1 woman being pregnant at time of exposure. Though national incidence of prenatal kratom use is unknown, 5 peer-reviewed case reports describe maternal and infant kratom withdrawal symptoms; two cases involved infants who were only exposed to kratom during the prenatal period, and both required treatment with a morphine weaning protocol to manage symptoms of neonatal abstinence syndrome.^11^ WPC also received 3 calls related to unintentional kratom ingestion in infants aged <2 years. As with any other psychoactive substance, public health messaging and clinical guidance to adults who use kratom should consider including information about safe storage practices to avoid unintentional ingestion or misuse by children.

As a final point, we consider the high prevalence of agitation among persons admitted to critical care to be worth noting. Again, extricating the role of kratom among these call data is challenging given small numbers in our dataset and the concomitant use of other substances in 5 of 7 critical care admissions. However, clinicians and toxicologists should recognize that although kratom does have sedative, opioid-like properties at higher doses, it also can act as a significant stimulant at lower doses,^2,3^ which is perhaps evidenced by prevalent agitation reported in WPC calls.

In conclusion, during January 1, 2010–September 1, 2022, in Wisconsin, kratom-associated exposure calls to the WPC have been increasing in frequency, were commonly reported as polysubstance exposures, and occasionally indicated intensive care unit admission. Continued research may help to more fully define kratom’s risk-benefit profile. Meanwhile, Wisconsin clinicians and public health experts can (i) be aware of its increasing prevalence, (ii) expand the collection of data specific to kratom use and exposure among patients—during the clinical documentation of patient history for example, and (iii) utilize available scientific literature to promote education materials for adults who choose to use kratom, particularly if they do so alongside other substances.

## Data Availability

All de-identified data produced in the present study are available upon reasonable request to the authors.

## Footnotes

† See e.g., 45 C.F.R. part 46, 21 C.F.R. part 56; 42 U.S.C. §241(d); 5 U.S.C. §552a; 44 U.S.C. §3501 et seq.

## Notes

### Competing Interest Statement

The authors have declared no competing interest.

### Funding Statement

This study did not receive any funding.

### Author Declarations

This activity was reviewed by the U.S. Centers for Disease Control and Prevention (CDC) and was conducted consistent with applicable federal law and CDC policy. See e.g., 45 C.F.R. part 46, 21 C.F.R. part 56; 42 U.S.C. section 241(d); 5 U.S.C. section 552a; 44 U.S.C. section 3501 et seq.

